# Online Mindfulness for Later Life: a feasibility study of a Public Mental Health intervention to increase resilience for Older Adults

**DOI:** 10.64898/2026.04.01.26349967

**Authors:** Adele Pacini, Naoko Kishita, Gemma Hawkins, Michelle Nicholson, Annabel Stickland, Rebecca Gould

**Affiliations:** Marie Curie Palliative Care Research Department, Division of Psychiatry, University College London,6th Floor, Maple House,149 Tottenham Court Road, London, W1T 7NF, UK; School of Health Sciences, University of East Anglia, Norwich, NR4 7TJ, UK; The Mindful Life Group, Farthingworth, Lynford, IP265HN, UK; Institute of Psychiatry, Psychology & Neuroscience (IoPPN), 16 De Crespigny Park, London SE5 8AF, UK; Division of Psychiatry, University College London, 6th Floor, Maple House,149 Tottenham Court Road, London, W1T 7NF UK

## Abstract

**Background:** Resilience is acknowledged to be an important component for successful aging in older adults, but there is scant evidence with which to inform public health interventions for this age group. The aim of this study is to determine whether the public health intervention, ‘mindfulness for later life’ is both feasible and acceptable to older adults.

**Methods:** Participants were recruited from September 2021 to June 2022 through older adult organisations and charities, such as the University of the Third Age, Age UK, and Age Concern, and by adverts distributed through village newsletters and support organisations. Participants were offered six weekly sessions of mindfulness therapy, the program was based on the mindfulness-based stress reduction program, each session was two hours long with 10-15 participants per program. The following two pre-defined indicators of success needed to be met for the program to be deemed feasible: successful uptake (recruitment of 30 participants over nine months) and initial engagement (≥70% completing at least four online sessions).

**Results:** Thirty-three potential participants were screened for eligibility over nine months, 31 of whom were recruited to the study (103% of the target sample). Of these, 28 participants (90%) completed four or more online sessions. Thus, predefined indicators of feasibility were met.

**Conclusions:** This study supports the feasibility of delivering the mindfulness for later life program as a public health intervention, including recruitment and treatment completion. A full-scale trial to assess the clinical- and cost-effectiveness of the intervention including its long-term effects is warranted.

## Introduction

Resilience is recognised as an important factor in successful ageing among older adults, yet there is little evidence to inform public mental health (PMH) interventions aimed at enhancing resilience in this demographic (Madsen et al., 2019). In older adults, common psychosocial stressors such as loneliness, social isolation, and financial struggles intersect with higher occurrences of bereavement, caregiving duties, physical ailments, and cognitive decline, leading to an elevated risk of mortality (Steptoe et al., 2013). This underscores the importance of PMH interventions in bolstering resilience in older adults to meet these challenges.

Recent systematic reviews and meta-analyses have highlighted resilience as a modifiable factor with significant potential to protect the overall health of older adults, particularly their mental well-being (Avila et al., 2016; Farber & Rosendahl, 2020). Resilience is the ability to handle, and adjust to, various forms of adversity, such as stress, trauma, abuse, and poverty. While it can be conceptualised at both the community and individual levels, this study specifically focuses on psychological resilience at the individual level. This type of resilience has been found to contribute to increased life satisfaction, happiness, hope, self-esteem, social support, longevity, and overall quality of life in older adults (Gallardo-Peralta et al., 2020; Lai et al., 2021; MacLeod et al., 2016).

Madsen et al. (2019) reviewed the current status of the evidence related to both community and individual resilience in community dwelling older adults. Of the eleven studies examining individual resilience, the majority were descriptive qualitative studies which sought to understand older adults’ perspectives on resilience. Only two of the studies, both of which were qualitative, explored changes after an intervention (Community Men’s Sheds, Moylan et al., 2015; Videoconference education groups, Banbury et al., 2017). In neither of these studies was resilience directly assessed through quantitative outcomes.

The evidence for PMH interventions to improve resilience, wellbeing and prevention of mental illness for older adults is similarly limited. In a recent systematic review, Kingstone et al. (2022) found that there is a ‘dearth of evidence’ on mental health interventions for older adults delivered by non-traditional providers. Similarly, a scoping review by Lee et al. (2021) noted that ‘more robust evidence is required’ on community-based interventions for the prevention of mental ill health and promotion of wellbeing in older adults in the UK. Of the studies reviewed by Lee et al., (2021) none addressed or measured resilience.

Of the psychological interventions known to improve resilience, mindfulness arguably has the strongest evidence base and is well placed to deliver benefits across the scope of PMH practice (for reviews see Joyce et al., 2018; Donaldson-Feilder, 2019). Approaches like mindfulness speak to the universal nature of both wellness and suffering and improvements in resilience have been reported across both physical (e.g. Kabat-Zinn, 1982) and mental health conditions (e.g. Segal et al., 2013). Several systematic reviews have suggested that mindfulness approaches may be cost-effective to foster individual resilience (Duarte et al., 2019; Joyce et al., 2018, Galente et al., 2021). For older adults, Kayser et al. (2022) report that the mindfulness evidence base is ‘promising’ for improving ability to cope in older adults with chronic health conditions.

Despite this, mindfulness has been surprisingly absent from discussion in public health fields (Oman, 2023). Of note, Galente et al.’s (2021) meta-analysis of mindfulness delivered in non-clinical settings across the adult age range demonstrated that mindfulness-based practices (MBPs) on average improved medium term mental health outcomes in nonclinical settings. The authors suggested that MBPs targeted at populations at higher risk of developing mental health difficulties, or those with subclinical mental health difficulties were more beneficial. There are potential further benefits for older adults, including enhanced attention control, preserved neural functioning and reduced systemic inflammation (Larouche et al., 2014; Fountain-Zaragoza and Prakash, 2017), all of which can help to stem cognitive decline (Livingston et al., 2020)

The current literature points to the potential benefits of adopting a public health approach and offering mindfulness for fostering resilience in older adults, as well as a clear need to build the research literature on PMH interventions for this population. We therefore aimed to determine whether it is feasible to deliver an online PMH mindfulness intervention to older adults in the UK, particularly in terms of recruitment and treatment completion, to inform a future definitive trial.

## Methods

### Reporting Guidelines

All reporting adheres to the Consolidated Standards of Reporting Trials (CONSORT) guidelines for transparent reporting of feasibility trials (Schulz et al., 2010), and the Template for Intervention Description and Replication (TIDieR) guidelines for detailing interventions (Hoffmann et al., 2014). This ensures comprehensive reporting of both trial procedures and the mindfulness intervention itself to facilitate replication.

### Design

This is a single-arm feasibility study of a community-based online Mindfulness for Later Life (MLL) program. Data were collected as part of a service development and evaluation program for the Gatehouse Charity in East Anglia. Participants gave informed consent for their data to be used for research purposes. Ethical approval for the secondary analysis of the data for research purposes was provided by the UCL Research Ethics Committee (ID 23849.001).

### Participants

Participants were eligible if they were aged 65 years and over, and could speak and comprehend English. Participants had to be able to commit to the full program and associated home practice, and live in Norfolk or Suffolk. All participants needed to have access to the internet either at home or in a quiet location where they would not be disturbed, and a device to access the videoconferencing platform. Exclusion criteria included a dementia diagnosis in the moderate range of severity (determined either by prior clinician diagnosis or an Addenbrooke’s Cognitive Examination score ≤78, (Mioshi et al., 2006)) active suicidal ideation, uncontrolled bipolar disorder or schizophrenia.

### Recruitment Settings

Participants were recruited from September 2021 to June 2022 through older adult organisations and charities, such as the University of the Third Age, Age UK, and Age Concern, and by adverts distributed through village newsletters and support organisations, over a nine-month period. The program was promoted as helping to improve quality of life and resilience as well as reduce day to day stresses, anxiety, depression, carer burnout and fatigue.

### Intervention

The MLL program was delivered synchronously online via the Zoom video conferencing platform. The program was based on the mindfulness-based stress reduction program (Kabat-Zinn, 1982) and ran weekly for six weeks with 10-15 participants per program. The content was adapted for Older Adults (eg by removing reference to ‘work stress’) and the reading age of both the practices and the handouts was reduced to age seven (Gunning Fog index) to be accessible for participants with mild cognitive difficulties. The program was delivered by a Chartered Clinical Psychologist and registered mindfulness teacher with the British Association of Mindfulness Based Approaches. As well as the program teacher, an assistant attended to help with any technological issues. Sessions ran for 2 hours, and guided practices were approximately 20 minutes long. See Supplementary material 1 for a detailed description of each session.

### Procedure

Participants self-referred, with both applications and consent forms submitted online via a survey platform, applications were accepted across the year, with programs starting three times per year. Application forms recorded key demographic information (as listed in Table 1), along with history of head injury, and current diagnoses of dementia and/or mental health diagnoses. Each program had a maximum of 15 participants; any participants who applied to a full program were advised that they would be allocated to the next available group. Two weeks before the program began participants completed the online psychometric questionnaires. They also met with the program facilitator via zoom videoconferencing platform, to ensure that the participant met the inclusion criteria, could become familiar with the facilitator, could ask any questions, and check that the video conferencing software was running smoothly. Any technological issues were passed to an assistant who resolved them with the participant before the program began.

### Feasibility outcomes Primary outcomes

The following two pre-defined indicators of success needed to be met for the program to be deemed feasible: successful uptake (recruitment of 30 participants over nine months) and initial engagement (≥70% completing at least four online sessions). These indicators were agreed with the funders (Norfolk Community Foundation and West Suffolk Integrated Care Board) before the program started.

### Secondary outcomes

The following data were also examined to inform the feasibility of the program: *Recruitment, eligibility and attrition*. The number of self-referrals, reasons for refusal, numbers ineligible, reasons for ineligibility and numbers lost to follow-up.

*Resulting sample characteristics*. Descriptive demographic data for all participants who attended the screening session and those who were eligible and invited to the intervention phase.

*Treatment completion and acceptability*. The number of sessions completed in the 6-week intervention phase, and reasons for withdrawal from the intervention.

### Psychometric outcomes

The outcome measures were taken at baseline and after the six-week program to examine the feasibility of psychometric outcomes data collection. The resilience measure used was the Brief Resilience Scale (BRS; Smith et al., 2015). The BRS is a short, self-report psychometric measure that assesses resilience as the ability to bounce back or recover from stress and adversity. Cut off scores as recommended by Smith et al. (2015) are: 1.00-2.99 = Low resilience; 3.00-4.30 = Normal resilience and 4.31-5.00 = High resilience. Additional psychometric measures assessed depression (Patient Health Questionnaire, PHQ-9; Manea et al., 2012), anxiety (Generalised Anxiety Disorder scale, GAD-7; Spitzer et al., 2006), attitudes and behaviours associated with mindfulness (Cognitive and affective mindfulness scale revised, CAMS-R; Feldman et al., 2007), quality of life (Older Persons Quality of Life scale, OPQOL; Bowling et al., 2009), and compassionate beliefs and behaviours towards oneself and others (Relational Compassion Scale, RCS; Hacker et al., 2008). The RCS was included as a measure of self and other-compassion, as compassion is both known to improve resilience (Bag et al., 2022), and has been shown to increase with mindfulness training (Tirch, 2010). The measures are validated and widely used; they are also appropriate for use with older adults, including those who may have some level of cognitive impairment.

### Sample size

A sample size of 30 is consistent with recommendations for sample sizes of 24-35 for feasibility studies when estimating the standard deviation for a continuous outcome (Teare et al., 2014; Whitehead et al., 2016).

### Statistical methods

The aim of a feasibility evaluation is not to assess the efficacy of an intervention and thus no formal statistical analysis was conducted, as recommended for pilot or feasibility studies (Lancaster et al., 2004; Thabane et al., 2010). Psychometric measures were summarised using means and standard deviations (SD). However, to identify signals of efficacy, effect sizes (Cohen’s *d*) were calculated for each outcome using cases for which both pre- and post-intervention data were available. These were calculated by subtracting the pre-group mean from the post-group mean and dividing by the pre-SD (Durlak, 2009).

## Results

### Primary outcomes

Thirty-three potential participants were screened for eligibility over nine months, 31 of whom were recruited to the study (103% of the target sample). Of these, 28 participants (90%) completed four or more online sessions. Thus, predefined indicators of feasibility were met.

### Secondary outcomes

The number of participants, reasons for refusal, number of sessions completed, and numbers lost to follow-up are presented in Figure 1. All 33 potential participants who applied were screened for exclusion criteria as outlined above, and all participants were found to be eligible. Of these participants, two of the 33 (6%) declined to attend the intervention. Three of the 31 participants (10%) did not complete post-intervention psychometric measures.

**Figure 1.**
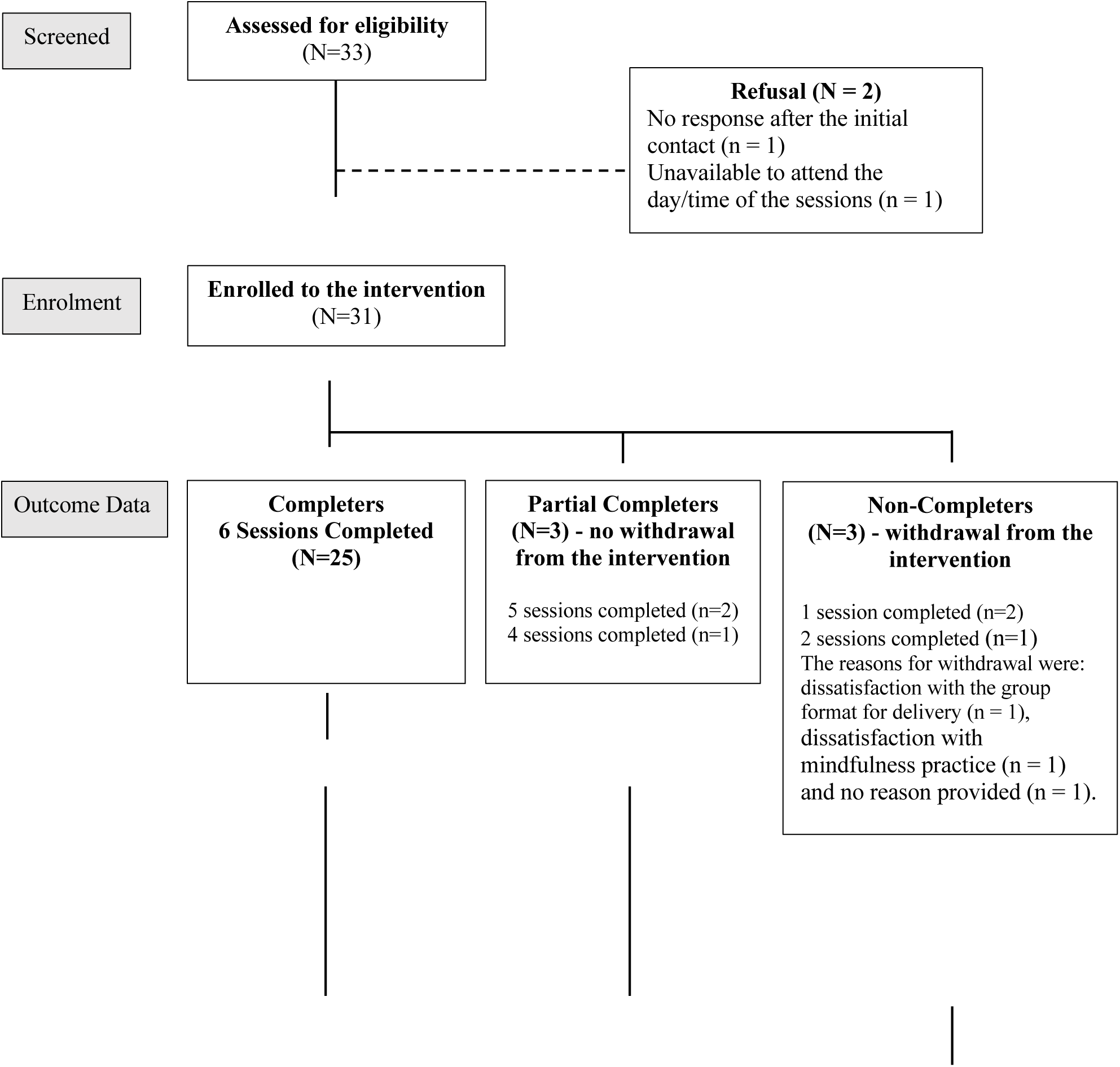
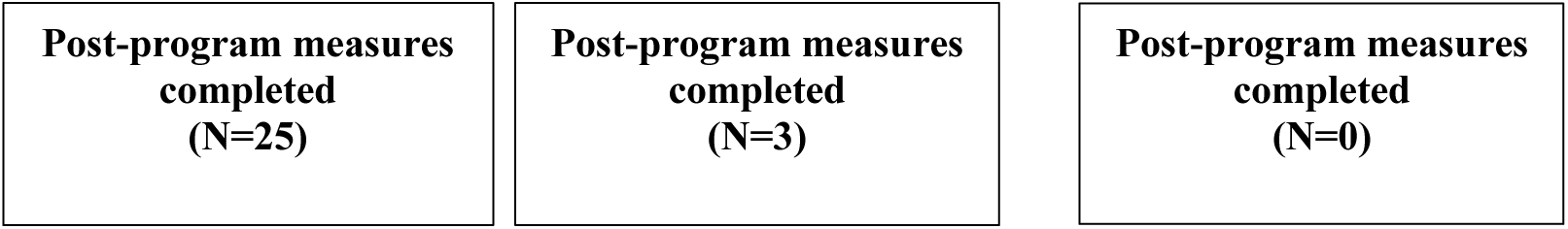
**MLL feasibility evaluation flow diagram.**

*See Figure 1. MLL feasibility evaluation flow diagram*.

### Resulting sample characteristics

Participant characteristics are summarised in Table 1. Of the 28 participants who completed the intervention (defined as completing four or more sessions), the majority of participants were female (82%), in the 70-79 years age range (53%) and lived in a rural setting (57%).

Participants’ resilience scores were in the low range on average at 2.83 (SD = 0.74). With regards to mental health, 50% of these participants were above the recommended cut-off of five on the PHQ-9 for depression, and 43% above the cut-off of five on the GAD-7 for anxiety.

*See Table 1 - Characteristics of participants at baseline/before the intervention*.

### Participant feasibility

Of the 31 participants, 25 (81%) completed all six sessions and 28 (90%) completed four or more sessions. Three participants (10%) withdrew from the intervention before the end of the program (see Figure 1). The reasons for withdrawal were: dissatisfaction with the group format for delivery (n = 1), dissatisfaction with mindfulness practice (n = 1) and no reason provided (n = 1).

### Psychometric Outcomes

Table 2 shows the means and standard deviations for all psychometric outcomes after completion of the program, and pre-post effect sizes (N=28). Changes in scores from pre to post intervention show that participants’ anxiety (GAD-7) and depression (PHQ-9) scores both decreased following the intervention. Further, participants, resilience (BRS), quality of life (OPQOL-Brief), mindfulness (CAMS-R) and relational compassion (RCS) all increased. *See Table 2 - Means and standard deviations for psychometric outcomes at follow-up and effect sizes (N=28)*

## Discussion

### Main finding of this study

Our results demonstrate that it is feasible to recruit participants and deliver online mindfulness therapy to older adults as a PMH intervention. We further showed that this intervention appeared to be acceptable to older adults, as indicated by recruitment and retention rates. The pre-defined indicators of success were met: 103% of the target sample was recruited over nine months and 90% completed at least four online sessions. Secondary outcomes showed that 81% of participants completed all six sessions, the attrition rate was low (10%) and data completion rates were high at 90%. Data were also suggestive descriptively of pre-post improvements in outcomes measuring resilience, depression, anxiety and relational compassion.

Our attrition rate of 10% is comparable to the 16% attrition reported in recent studies on online mindfulness delivered to older adults (Galuluzzi et al., 2024), and is lower than the attrition range of 35-92% from a systematic review of online mindfulness interventions for adults aged 18 upwards (Sommers-Spijkerman et al., 2021).

All participants self-referred from the community in this study, which provides important evidence regarding the potential of a PMH approach to offering psychological intervention to a population that is currently underserved by statutory services. Demographic data of those completing the intervention show that participants’ resilience scores prior to starting the intervention were, on average, in the low range, which improved to the normal range on completion of the program. Half of the participants had some symptoms of depression at pre-intervention (PHQ-9 score above five), with 18% scoring ten or more on the PHQ-9, indicating major depressive disorder (Manea et al., 2012). For anxiety, 43% of participants had some symptoms of anxiety at pre-intervention, with 18% scoring above 10 on the GAD-7, consistent with Generalised Anxiety Disorder (Spitzer et al.,2006). This is consistent with the estimated 37-43% of older adults reporting some symptoms of depression or anxiety (Braam et al., 2014; Rodda et al., 2011).

### What is already known on this topic

Despite significant rates of both depression and anxiety in this population, fewer than one in every six older adults seek medical attention for their symptoms (Burns, 2022). Ageism, a lack of mental health awareness, and a refusal to disclose symptoms are all reported as barriers to seeking help (Farrer et al., 2008; Pywell et al., 2020). From a public health perspective, untreated mental health difficulties in older adults are not simply an issue of equity of access. Improvements in both depression and anxiety following psychological therapy are associated with a reduced risk of dementia in older adults (John et al., 2022; Stott et al., 2022).

### What this study adds

Although this study was not powered to examine effectiveness, there was some preliminary evidence suggestive of improvements in scores for resilience, relational compassion, anxiety and depression (i.e., signals of efficacy). Encouragingly, effect sizes were largest for resilience and relational compassion, aligning with the PMH focus of the intervention. The relational compassion measure was included to capture the effect of mindfulness training on compassion (e.g. Neff & Dahm, 2015; Tirch, 2010); a component of our program is specific training in compassion and loving kindness for self and others. The precise relationships between resilience and self-compassion are still being unravelled (see Bag et al., 2022) but in older adults, compassion has been shown to improve resilience (Moore et al., 2015; Smith, 2015), coping ability and adaptation to stressful situations (Perez-Blasco et al., 2016; Tavares et al., 2023). Improvements in compassion and resilience levels observed in our study are very promising with respect to the potential for mindfulness to support the mental health of older adults in the community.

### Limitations of this study

There were some methodological limitations, given that this was a feasibility study, it was not adequately powered to examine the effectiveness of the intervention, and findings are therefore reported descriptively. Participant recruitment took place in two counties in the East of England. The population of the county that had the largest number of recruits is more than 90% White British, thus, the ethnic diversity of the sample was limited. This was a secondary analysis of a service evaluation data set that did not include a control group.

Therefore, preliminary evidence of improvements in outcomes should be interpreted with caution. The recruitment was to a community-based program rather than to a research study and this may overestimate recruitment and retention in a research trial. The intervention was delivered by an experienced clinician and mindfulness teacher, and the study did not test the feasibility of teaching other staff to deliver the program. There was no formal testing of treatment fidelity or follow-up after the intervention, it is therefore unclear whether any beneficial gains were sustained after intervention completion.

## Conclusion

This study provides evidence for the feasibility and acceptability of online mindfulness for older adults delivered as a public mental health intervention. There were some indications of improvements in resilience, relational compassion, anxiety and depression. However, this was a single-arm underpowered feasibility study and as such, these results should be interpreted with caution. It will be important to ascertain whether it is both feasible and acceptable to recruit older adults to a research trial of this intervention, ahead of a full-scale RCT to assess the clinical and cost effectiveness of the intervention including it’s long term effects.

## Data Availability

All data produced in the present study are available upon reasonable request to the authors

---

**Table 3.**
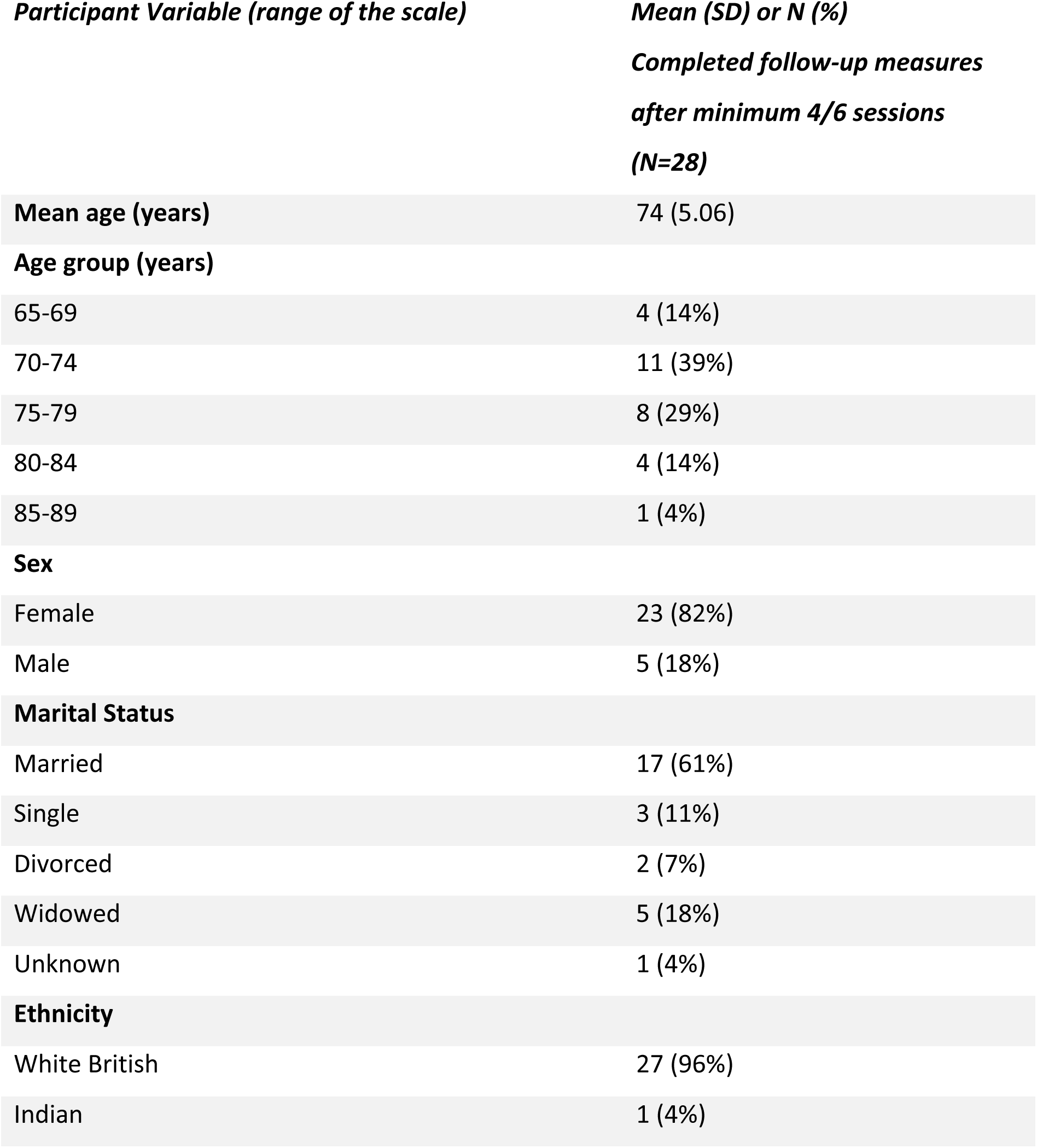
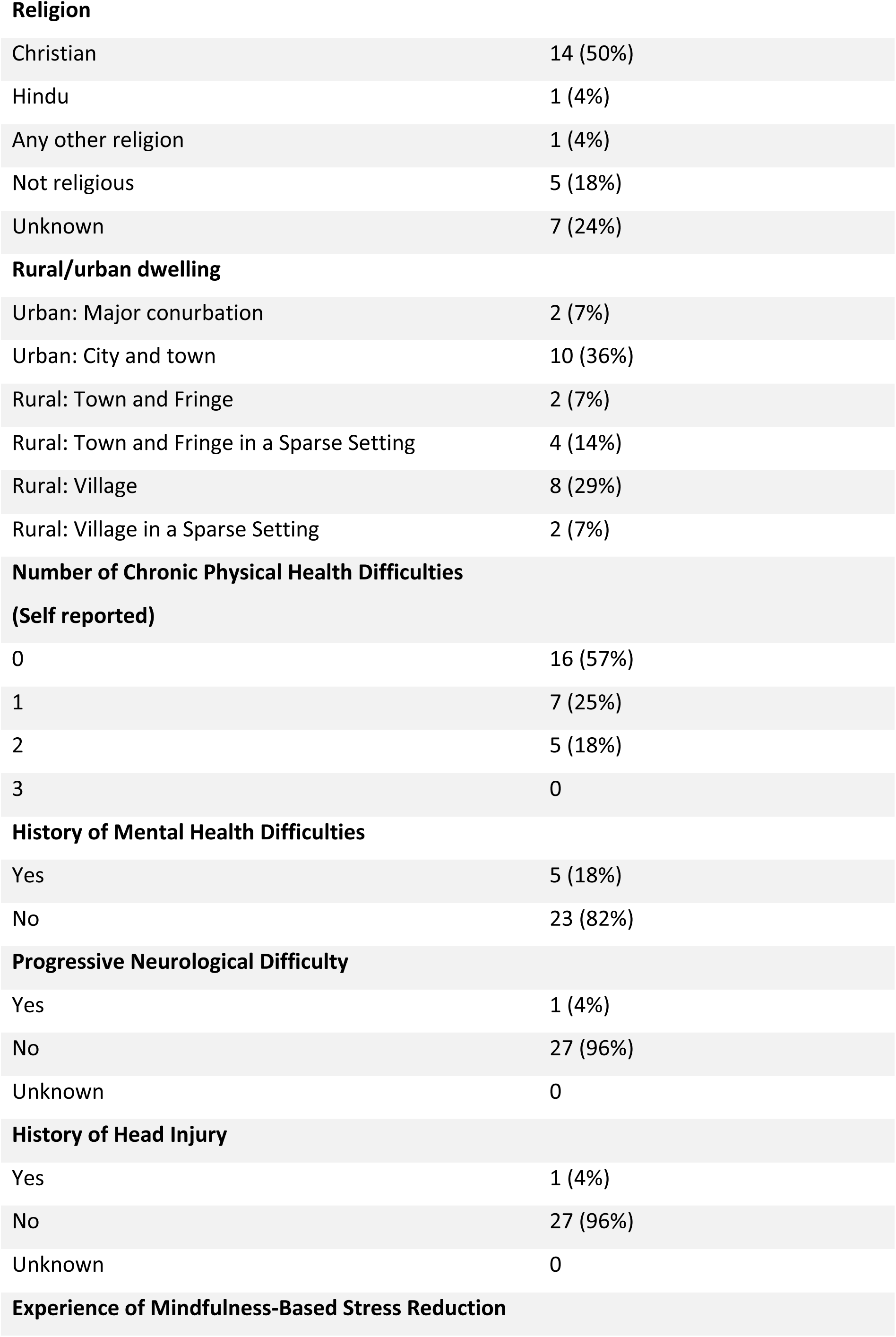
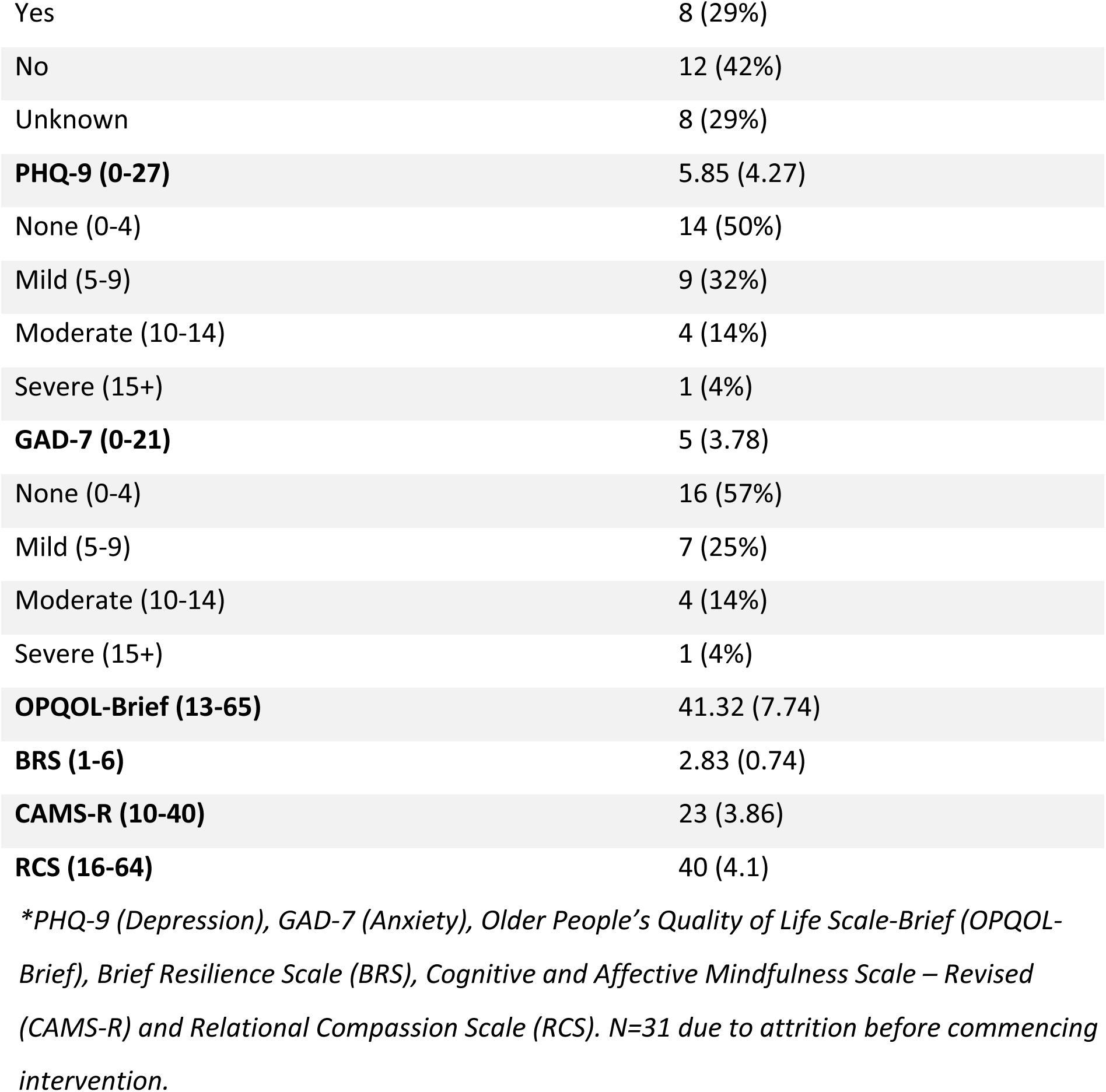
Characteristics of participants at baseline/before the intervention.

**Table 4.**
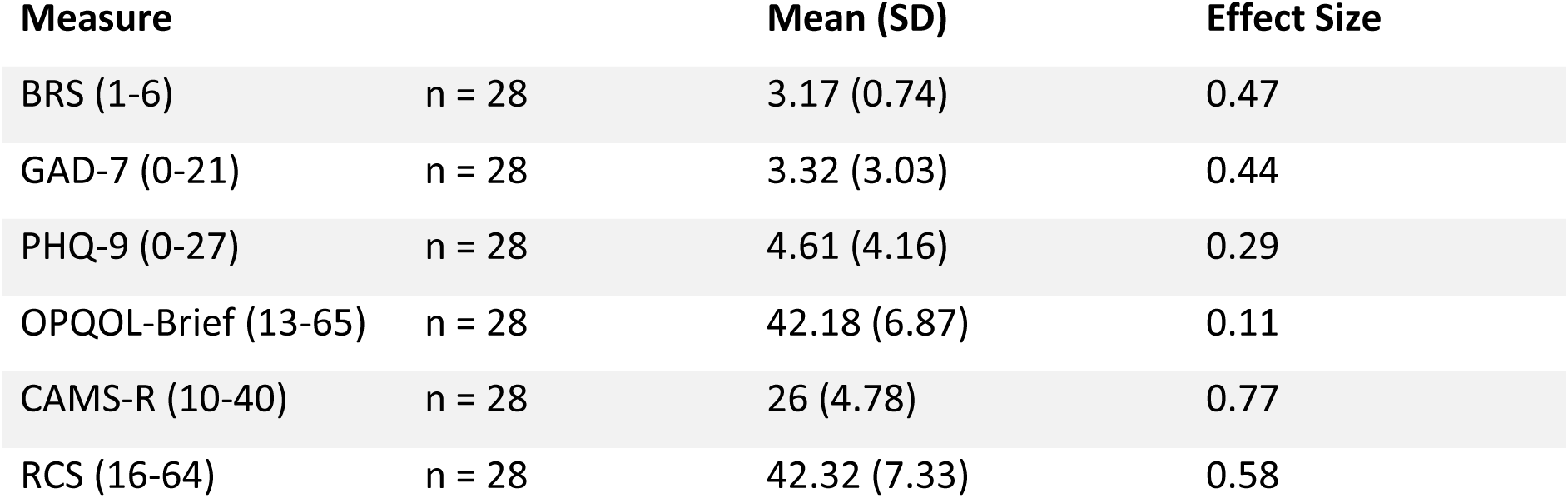
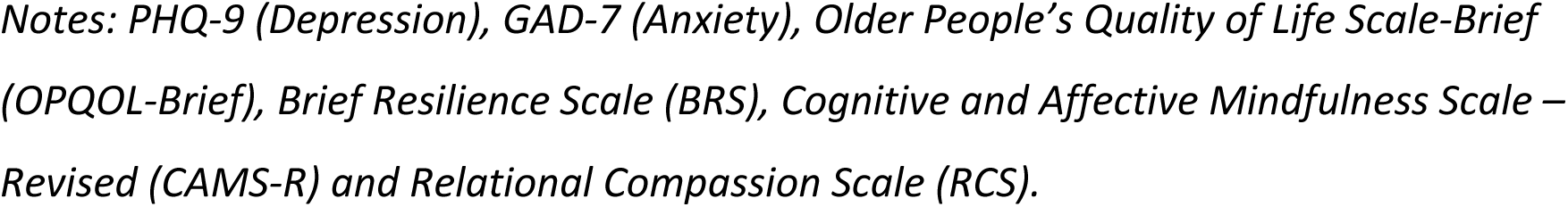
Means and standard deviations for psychometric outcomes at follow-up and effect sizes (N=28)

| Measure |  | Mean (SD) | Effect Size |
| --- | --- | --- | --- |
| BRS (1-6) | n = 28 | 3.17 (0.74) | 0.47 |
| GAD-7 (0-21) | n = 28 | 3.32 (3.03) | 0.44 |
| PHQ-9 (0-27) | n = 28 | 4.61 (4.16) | 0.29 |
| OPQOL-Brief (13-65) | n = 28 | 42.18 (6.87) | 0.11 |
| CAMS-R (10-40) | n = 28 | 26 (4.78) | 0.77 |
| RCS (16-64) | n = 28 | 42.32 (7.33) | 0.58 |

*Notes: PHQ-9 (Depression), GAD-7 (Anxiety), Older People's Quality of Life Scale-Brief (OPQOL-Brief), Brief Resilience Scale (BRS), Cognitive and Affective Mindfulness Scale – Revised (CAMS-R) and Relational Compassion Scale (RCS).*

**Supplementary material 1:** The main themes, activities and associated home practices for each week of the MLL Group.

| Week | Theme | Activities | Home practice |
| --- | --- | --- | --- |
| 1 | Automatic Pilot -<br>Mindfulness starts when we recognise the tendency to live on ***automatic pilot*** and commit to learning how best to step out of it, to become aware of each moment. | Establish mindful orientation of classes:<br>Raisin Exercise<br>Body Scan exercise and inquiry<br>Small group work – introductions to each other | Body Scan x 6 days<br>1 daily mindful activity |
| 2 | Working with barriers – understanding what practical and emotional barriers might get in the way of our daily practice. | Body Scan<br>Exhaustion funnel<br>Small group work – barriers and solutions<br>3 step breathing space | Body Scan and 3 step breathing space x 6 days<br>Daily mindful routine activity |
| 3 | Mindfulness and stress – “It’s not the stressors, but how you handle them” which influences the effects they have on health in mind and body. | 3 step breathing space<br>Primary and secondary stressors<br>Small group work – how do you respond to stress.<br>Mindfulness of breathing | Mindfulness of breathing x6 3 step breathing space x6 informal practice bringing awareness to a stressful activity. |
| 4 | Responding to thoughts – thoughts as mental | Mindfulness of breathing | Mindfulness of sounds and |
|  | events, rather than facts.<br>Starting to notice our thinking patterns, learning to observe them rather than becoming 'tangled' in them | Examples of Unhelpful thinking patterns<br>Small group work – what unhelpful thinking patterns are you aware of?<br>Mindfulness of sounds and thoughts | thoughts/ informal – bringing awareness to a pleasant activity. |
| 5 | Compassion – mindfulness is not just about awareness but also *how* we are aware.<br>Emphasising the kindly and compassionate attitude to our experiences | Mindfulness of sounds and thoughts<br>Mindful attitudes to our experiences<br>Small group work – are we kind to ourselves?<br>Mindful movement | Practice – Mindful movement,<br>Informal – bringing a compassionate attitude to everyday activities. |
| 6 | Keeping your practice alive. What do I need to do to take care of myself?<br>What intention to I want to set for myself? | Mindful movement<br>Listing an 'average' day – which activities are nourishing, which are depleting?<br>Small group work – discussion on the activities exercise above.<br>Body scan | Encouraged to continue to practice every day. Regularity rather than duration is important |

